# Proteomic analysis identifies lipoprotein(a)-associated proteins linked to incident atherosclerotic cardiovascular disease events

**DOI:** 10.64898/2025.12.16.25342337

**Authors:** Tiffany R. Bellomo, Seyedmohammad Saadatagah, Jiwoo Lee, Emily E. Bramel, Layla Abushamat, Anika Misra, Tetsushi Nakao, Satoshi Koyama, Aniruddh P. Patel, Sarah Urbut, Christie MP Ballantyne, Pradeep Natarajan

## Abstract

**Background:** The pathways linking lipoprotein(a) (Lp[a]) to atherosclerotic cardiovascular disease (ASCVD) are unclear. This study aimed to discover Lp(a)-associated plasma proteins and estimate their associations with incident ASCVD.

**Methods:** We analyzed 48,859 UK Biobank participants with measured Lp(a) and proteomic profiles, with replication in 9,416 individuals in the Atherosclerosis Risk in Communities (ARIC) study cohort utilizing a separate proteomic platform. Linear models assessed associations between Lp(a) and protein concentrations adjusted for age, sex, cigarette smoking, diabetes diagnosis, body mass index, systolic blood pressure, hypertension, low-density lipoprotein cholesterol (LDL-C), high-density lipoprotein cholesterol, triglycerides, estimated glomerular filtration rate, statin prescription, and the first 10 components of genetic ancestry. Multiple testing correction was performed using the Benjamini-Hochberg FDR method (P < 0.05). We examined how the protein effect sizes from the primary analysis using the outcome of Lp(a) aligned with those for the outcomes of an LPA genetic risk score (GRS) and LDL-C. Cox proportional hazards models quantified hazard ratios (HRs) for protein associations with incident ASCVD.

**Results:** Participants were a mean age of 57 years (SD 8.22), 93.9% European, and 53.8% male, with median follow-up of 8.9 years (IQR 8.3–9.7). Of 1,459 circulating proteins, 164 were significantly associated with Lp(a) after FDR correction, with enrichment for lipid degradation, metabolism, and insulin secretion. In the ARIC study, 10 proteins were replicated with consistent effect estimates. Of these replicated proteins, there were no significant associations observed with an *LPA* GRS. Only REG4 and VWC2 showed concordant associations with LDL-C (P < 0.001), consistent with their association with Lp(a). Five proteins exhibited concordant associations with Lp(a) and incident ASCVD (ITIH3, DLL1, REG4, VWC2, CBLN4). ITIH3 was positively associated with coronary artery disease (HR 1.13, 95% CI 1.04–1.23), peripheral artery disease (HR 1.42, 95% CI 1.19–1.69), major adverse limb events (HR 1.65, 95% CI 1.14–2.40), carotid stenosis (HR 1.45, 95% CI 1.13–1.85), and ischemic stroke (HR 1.33, 95% CI 1.13–1.55). CBLN4 uniquely showed inverse associations with Lp(a) and disease: higher levels were linked to lower risk of CAD (HR 0.88, 95% CI 0.80–0.96), PAD (HR 0.78, 95% CI 0.64–0.96), and ischemic stroke (HR 0.72, 95% CI 0.60–0.85).

**Conclusion:** Using high-throughput proteomics, we discovered and replicated 10 proteins associated with circulating Lp(a), several of which were independent of genetically-predicted Lp(a). While Lp(a) is highly heritable, these atherogenic proteins represent a non-heritable Lp(a) axis.

**Clinical Perspective:** *What’s New?:*

- Ten proteins associated with circulating lipoprotein(a) levels were identified and independently replicated in an external cohort, with associations independent of a genetic risk score.
- Five proteins (ITIH3, CBLN4, FLL1, REG4, VWC2) were concordantly associated with lipoprotein(a) and incident atherosclerotic cardiovascular disease.

*Clinical Implications:*

- These proteins may represent pathways through which lipoprotein(a) drives atherosclerosis beyond traditional lipid mechanisms.
- Future mechanistic studies should investigate the roles of these proteins to guide the development of targeted strategies for preventing atherosclerotic cardiovascular disease.

## INTRODUCTION

Atherosclerotic disease remains the leading cause of morbidity and mortality in both men and women in the United States.^1^ Major adverse cardiovascular events continue to occur despite adherence to contemporary guidelines for lipid-lowering, blood pressure control, and glucose management,^2,3,4^ as well as efforts to optimize cardiovascular health. Lipoprotein(a) (Lp[a]) has emerged as a novel, potentially causal independent risk factor for coronary artery disease (CAD).^5^ Lp(a)-lowering therapies are currently being evaluated in clinical trials, and these efforts lay the groundwork for additional therapeutic development.^6,7^

Despite detailed characterization of Lp(a)’s structure and epidemiologic link with atherosclerotic disease, the precise cellular and molecular pathways that mediate its proatherogenic and prothrombotic effects remain unresolved. Lp(a) is a lipoprotein in which apolipoprotein(a) [apo(a)] is covalently linked to apolipoprotein B 100, which carries cholesterol, triglycerides, and oxidized phospholipids.^8^ Plasma Lp(a) concentrations are largely genetically determined, in which variation at the *LPA* locus accounts for most inter-individual variability.^9^ Beyond the contribution of its individual lipid components, the unique protein architecture of Lp(a) has been implicated in proatherogenic, proinflammatory, and antifibrinolytic mechanisms that promote plaque formation, inflammation, and calcification.^10^ Specifically, the accumulation of Lp(a) in the arterial wall leads to migration of activated monocytes and results in inflammation, atherosclerotic plaque development, and destabilization.^10^ However, the assembly, clearance, and receptor interactions of Lp(a), and how these processes translate to downstream vascular effects, are not fully understood.^11^

In this study, we leveraged UK Biobank (UKB) data to quantify the relationship between circulating Lp(a) and proteomic profiles, and to investigate the association of these proteins with incident atherosclerotic cardiovascular disease (ASCVD). In primary analyses, we used multivariable-adjusted linear models to test the association of Lp(a) with 1,459 circulating proteins and annotated significant proteins for their molecular functions. Proteins whose associations were replicated in the Atherosclerosis Risk in Communities (ARIC) study were carried forward for downstream analyses. We stratified participants by demographic groups to assess for effect modification. We then tested whether an Lp(a) genetic risk score (GRS) was associated with these validated proteins and evaluated whether observed associations were mediated by low-density lipoprotein cholesterol (LDL-C). Finally, we examined the relationships between these final candidate proteins and incident ASCVD events in the UKB.

## METHODS

### Design and population

This cohort study used data from the UKB, a population-based prospective cohort. Study baseline was defined as enrollment between 2006 and 2010, when blood samples for Lp(a) measurement^12,13^ and proteomic profiling^14,15^ were collected on the same day.^16^ Participants were followed prospectively through the most recent UKB data update available in 2024 (Supplemental Figure 1). Analyses were performed under UKB application number 7089, and all participants provided informed consent. The Mass General Brigham Human Research Committee Institutional Review Board approved the secondary analyses (IRB # 2021P002228).

The UKB is comprised of genetic and clinical data on over 500,000 individuals aged 40–69 years at recruitment (Supplemental Figure 1).^17^ Circulating Lp(a) was measured at baseline in 460,544 participants. For the present study, we restricted analyses to participants with baseline proteomic measurements to-date, yielding 52,705 baseline proteomic profiles using the Olink Explore 1536 platform. Baseline assessments, conducted at 22 centers across the UK, included medical history, lifestyle exposures, physical examination, and collection of biological samples. Clinical outcomes were ascertained through linkage to National Health Service records and UK death registry data, using ICD 9/ICD 10 codes, OPCS 3/OPCS 4 procedure codes, and death records. Participants were also excluded if they were missing demographic data (age, self reported race/ethnicity, or genetic ancestry), yielding a final cohort of 48,859 participants (Supplemental Figure 1).

### Lp(a) measurement as the primary exposure

Serum Lp(a) concentrations were measured as previously described by the UKB.^12^ Briefly, an immunoturbidimetric assay (Randox Laboratories, Crumlin, County Antrim, UK) using the Denka Seiken method was performed on a Beckman Coulter AU5800 platform; the assay’s clinical reportable range is 3.8–189 nmol/L. Values above 189 nmol/L (n = 32,953) were re-analyzed after serial dilution to obtain quantitative results. Lp(a) values were reported in nmol/L and were converted to mg/dL for some analyses using the factor 2.15 (nmol/L ÷ 2.15 = mg/dL).^18^ Measured lipid values for participants reporting lipid lowering therapy were adjusted to estimate untreated LDL-C C and HDL C, as previously described.

### Protein measurements as the primary outcome

Proteomic data were collected from participants in the UKB PPP as previously described.^14^ Non-fasting baseline plasma samples from 52,749 UKB participants were profiled on the Olink Explore 1536 platform using Olink Proximity Extension Assay (PEA) technology. Samples or proteins were excluded for high missingness (>10% missing protein measurements) or for excess relatedness (second degree relatives or closer). Remaining missing protein values in the quality controlled Olink dataset were imputed with the miceforest Python package version 6.0.0 using 10 fold cross validation and covariates of protein batch, study center, first 10 principal components of genetic ancestry, and all other protein values. During imputation using all other available protein values, we evaluated potential sources of technical and population structure bias by incorporating protein assay batch, study center, and the first 10 principal components of genetic ancestry. Finally, protein values were log2 transformed and standardized (mean = 0, SD = 1) to allow for comparison across analytes.

### Demographics and comorbidity ascertainment

Other standard cardiovascular risk factors used in this analysis were selected based on previously described cardiovascular risk prediction models,^19^ including age at enrollment, sex, cigarette smoking, diabetes diagnosis, body mass index (BMI), systolic blood pressure (SBP), hypertensive (HTN) medication, low-density lipoprotein (LDL-C) cholesterol, high-density lipoprotein cholesterol (HDL-C), triglycerides, estimated glomerular filtration rate (eGFR), statin prescription, and the first 10 components of genetic ancestry. Age at UKB enrollment was utilized in the analysis. Sex was derived from fixed categories of male and female. Current cigarette smoking was defined as lifetime smoking of at least 100 cigarettes and current smoking. Diabetes mellitus was defined as glycated A1c ≥6.5% or prior physician diagnosis.^20^ BMI was measured using Tanita BC-418MA body composition analyzer. Blood pressure was measured by the UKB staff at the time of enrollment. HTN medication was defined as prescription of an anti-hypertensive medication. All laboratory values for total and HDL-C were derived from a non-fasting sample collection assayed within 24 hours. eGFR was calculated using the Chronic Kidney Disease Epidemiology Collaboration equation.^21^ Statin prescription was defined as a prescription written at the time of enrollment. Sub-analyses were performed using both demographic data and ancestry. Ancestry was calculated using a k-nearest neighbors (KNN) algorithm based on genetic similarities.^22^

### Replication in ARIC

Significant proteins identified in primary analyses were carried forward for replication in the ARIC study, which is a large population-based cohort study in geographically diverse regions recruited from 1987 through 1989.^23^ This replication was performed using data from 11,637 participants with measured Lp(a) at visit 4 (1996–1998). Since Lp(a) levels are genetically determined and remain stable at least over several years,^24^ we used Lp(a) measurements from visit 4 (1996-1998) with proteomic measurements from the earlier visit 2 (1990-1992), which allowed for the longest possible follow-up period for incident cardiovascular outcomes.^25^

Protein measurements were performed using the SOMAscan v4 multiplexed SOMAmer assay (SomaLogic) as previously described.^26^ Briefly, plasma collected at ARIC visit 2 was stored at −80 °C and sent to SomaLogic for quantification. The assay uses modified single stranded DNA aptamers to bind proteins, with DNA detection to infer relative protein concentrations. ARIC data were combined with healthy control data and normalized prior to analysis, and SOMAmer measures were log2 transformed to reduce skewness. Initial data comprised of 5,284 aptamers. Quality control procedures included flagging samples if any of four sample calibration factors (hybridization control normalization factor and normalization factors for dilution factors 0.005%, 0.5%, and 20%) fell outside 0.4–2.5, and flagging SOMAmers if any intraplate calibration factor fell outside 0.8–1.2. The median coefficient of variation by the Bland–Altman method (CVBA) was 6.3%, and the median split sample reliability coefficient (intraclass correlation) for visit 2 QC was 0.93. Aptamers were excluded if CVBA >50% or variance <0.01 on the log scale, and aptamers binding mouse Fc fusion proteins (n = 228), contaminants (n = 15), or non protein targets/spuriomers (n = 70) were removed; six UniProt IDs and three protein names were manually annotated. Outlying sample values (>5 SD from the log2 mean for a given SOMAmer) were winsorized. After standard ARIC QC exclusions, 4,955 SOMAmers mapping to 4,712 unique proteins remained for analysis. Of the 164 proteins significantly associated with Lp(a) in the UKB primary analysis, 37 proteins were also measured in the ARIC study and carried forward for replication.

### Primary proteomic association analyses

Linear models were fit to examine the associations between Lp(a) levels (per 50 nmol/L increment) and protein concentrations in the UKB cohort. All models were adjusted for age, sex, cigarette smoking, diabetes diagnosis, BMI, SBP, HTN medications, LDL-C, HDL-C, triglycerides, eGFR, statin prescription, and the first 10 components of genetic ancestry. Statistical significance was defined as P < 0.05 after correction for multiple testing using the Benjamini-Hochberg false discovery rate (FDR) method. Effect sizes are reported as standardized beta coefficients with 95% confidence intervals. Proteins significantly associated with Lp(a) levels were functionally annotated using Gene Ontology (GO),^27^ UniProt (UP),^28^ and Reactome databases.^29^ Pathway enrichment analysis was performed using the Database for Annotation, Visualization, and Integrated Discovery (DAVID) tool version 6.8, with all of the Olink proteins assessed in the discovery analysis serving as the background reference.^30^ Enrichment was considered significant at FDR-adjusted P < 0.05. Significantly associated proteins were carried forward for replication in the ARIC study with SomaScan using identical statistical models and covariate adjustments. Pre-specified subgroup analyses were conducted stratified by sex and genetic ancestry.

### Secondary analysis

We compared the proteomic effects of measured Lp(a) levels with those of a clinically available LPA GRS and LDL-C cholesterol levels using standardized effect sizes.^31^ Kaplan-Meier survival curves and cumulative incidence plots were generated to visualize the relationship between protein quartiles and time to the first ASCVD event. Cox proportional hazards regression models were used to quantify hazard ratios (HR) with 95% confidence intervals for the association between protein levels and incident ASCVD including CAD, peripheral arterial disease (PAD), major adverse limb event (MALE), carotid stenosis, and ischemic stroke defined previously in the UKB.^32^ The age-specific HR for risk of atherosclerosis was plotted for each protein at each age of enrollment to understand time-varying effects.^33^ All statistical analyses were performed using R version 4.2.2 (R Foundation for Statistical Computing, Vienna, Austria. URL: https://www.R-project.org/).

## RESULTS

### Cohort

Primary analysis included 48,859 participants with proteomic profiles and measured Lp(a) concentrations, with a mean value of 56.25 (SD 76.03) nmol/L (Table 1). UKB participants were on average 57 (SD 8.22) years old at enrollment, 93.9% of participants were European, and 53.8% of participants were male, followed over a median period of 8.9 years (IQR 8.3–9.7) after the baseline blood draw. The age and sex distribution were similar in UKB and the ARIC study, but ARIC comprised a higher prevalence of non-European ancestry. In terms of comorbidities in UKB compared to the ARIC study, there was a lower prevalence of obesity (UKB 24.6% vs ARIC 27.6%), current smokers (UKB 10.6% vs ARIC 19.2%), and diabetes (UKB 6.0% vs ARIC 12.5%). There was a higher proportion of individuals on a statin medication in the UKB (UKB 17.5% vs ARIC 2.5%), but lipid profiles were similar between cohorts. CAD was more prevalent in the ARIC study (4.9%) than in UKB (2.5%).

**Table 1.** Summary of baseline characteristics of individuals studied from the discovery cohort UK Biobank and replication cohort Atherosclerosis Risk in Communities. Abbreviations: eGFR, estimated Glomerular Filtration Rate; LDL-C, Low-density Lipoprotein Cholesterol; HDL-C, High-density Lipoprotein Cholesterol; UKB, UK Biobank; ARIC, Atherosclerosis Risk in Communities.

| **Variable** | **UKB Participants**<br>**(n=48,859)** | **ARIC Participants**<br>**(n=9,416)** |
| --- | --- | --- |
| Age (years, mean (SD)) | 57.29 (8.22) | 56.90 (5.68) |
| Female Sex (%) | 22586 (46.2%) | 3844 (44.0%) |
| Ancestry (%) |  |  |
| African (%) | 1327 (2.7%) | 1716 (19.6%) |
| Admixed Americans (%) | 342 (0.7%) | 0 (0%) |
| Eastern Asian (%) | 231 (0.5%) | 0 (0%) |
| European (%) | 45896 (93.9%) | 7018 (80.4%) |
| South Asian (%) | 1063 (2.2%) | 0 (0%) |
| Obesity (%) | 11939 (24.6%) | 2411 (27.6%) |
| Current Smoker (%) | 5171 (10.6%) | 1678 (19.2%) |
| Systolic blood pressure (mmHg, mean [SD]) | 139.64 (19.72) | 120.37 (18.00) |
| Hypertension defined by medications (%) | 15038 (30.8%) | 2888 (33.2%) |
| Diabetes (%) | 2941 (6.0%) | 1092 (12.5%) |
| eGFR (mean [SD]) | 87.23 (16.97) | 74.72 (19.23) |
| Statin medication at enrollment (%) | 8550 (17.5%) | 215 (2.5%) |
| LDL-C cholesterol (mg/dL, mean [SD]) | 136.42 (34.06) | 132.77 (35.91) |
| HDL cholesterol (mg/dL, mean [SD]) | 55.79 (14.86) | 50.32 (16.56) |
| Triglycerides (mg/dL, mean [SD]) | 154.63 (90.43) | 128.43 (64.39) |
| Lp(a) (nmol/L, mean [SD]) | 56.25 (76.03) | 25.68 (29.93) |
| Coronary artery disease (%) | 12302 (2.5%) | 426 (4.9%) |

### Primary analyses

When testing the associations of the exposure Lp(a) per 50 nmol/L with the outcomes of 1,459 circulating proteins in UKB, 164 significant candidate proteins were identified (Figure 1, Supplemental Table 1). Enrichment analyses across multiple annotation schemes supported the biological plausibility of these proteins in atherosclerosis and Lp(a) mechanisms, including GO biological process annotations related to lipid degradation and metabolism, as well as UP annotations for feeding behavior and insulin secretion (Figure 1B-G). This was consistent with significant tissue enrichment in the pancreas and molecular functions related to growth factor binding, lysosomal localization, and galactoside-binding activity (Supplemental Figure 2).

**Figure 1.**
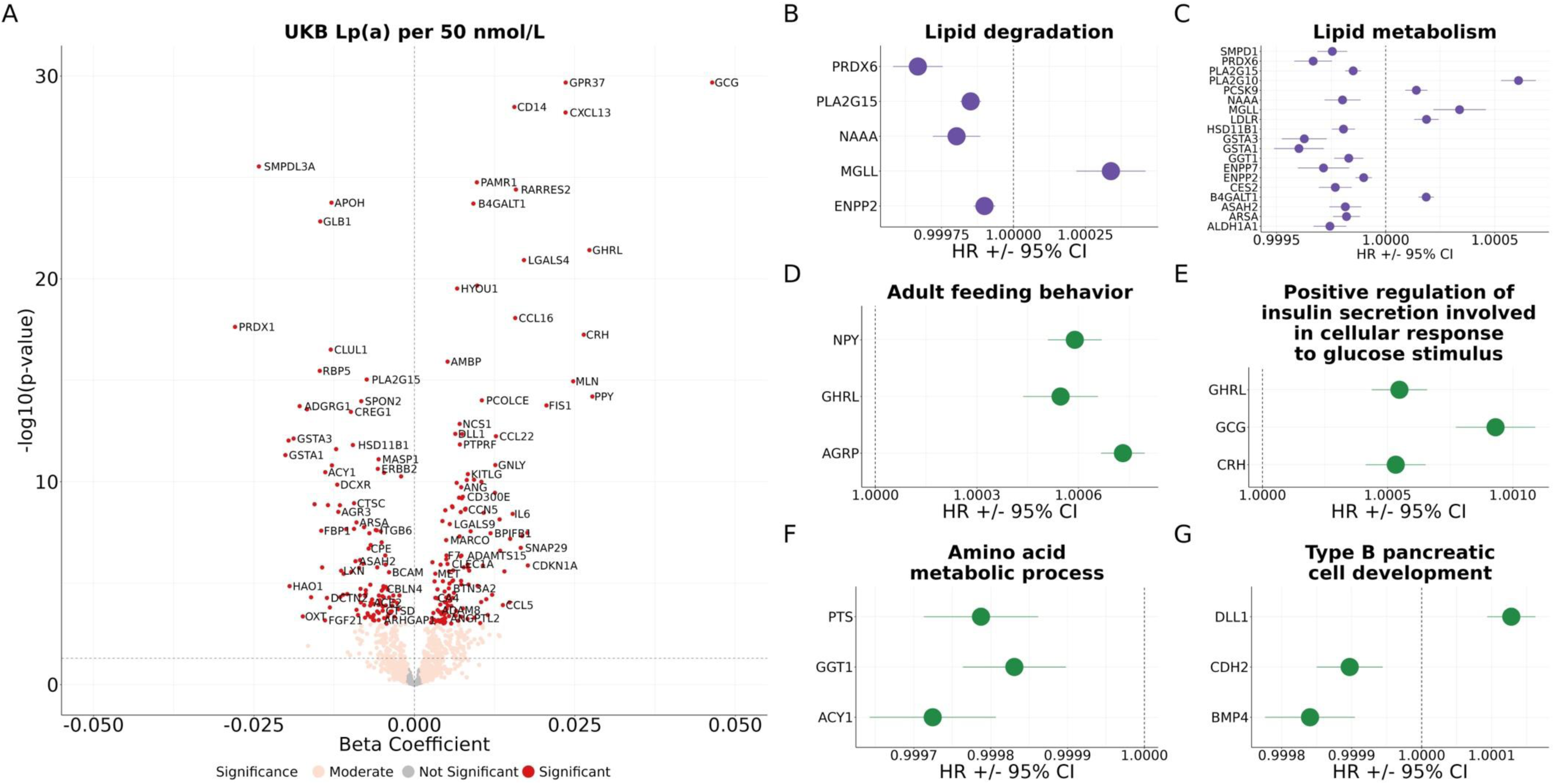
Association of Lp(a) per 50 nmol/L with circulating protein levels. (A) Volcano plot visualizing associations between all proteins and Lp(a) per 50 nmol/L, plotting −log10(p) against the corresponding β coefficient. (B) Forest plots of effect estimates and 95% CIs for proteins significantly associated with Lp(a) per 50 nmol/L. Purple indicates GO annotations, and green indicates UniProt annotations. Models were adjusted for age at enrollment, sex, smoking, DM, BMI > 30, SBP, eGFR, LDL, HDL, TG, statin use, and PC1–PC10. Multiple testing across 1,463 proteins was addressed using the BH FDR correction, with FDR-adjusted p < 0.05 considered significant. Abbreviations: BMI, body mass index; CI, confidence interval; DM, diabetes mellitus; eGFR, estimated glomerular filtration rate; FDR, false discovery rate; GO, Gene Ontology; HDL, high-density lipoprotein cholesterol; LDL, low-density lipoprotein cholesterol; Lp(a), lipoprotein(a); PC, principal component; SBP, systolic blood pressure; TG, triglycerides.

Of the 164 significantly associated proteins identified in the UKB using Olink, 10 proteins were replicated in the ARIC study using SomaScan with consistent effect estimates (Figure 2, Supplemental Table 2). Significant inverse replicated associations were observed for anterior gradient protein 3 (AGR3), cerebellin 4 precursor (CBLN4), cellular repressor of E1A-stimulated genes 1 (CREG1), dicarbonyl/L-xylulose reductase (DCXR), and ectonucleotide pyrophosphatase/phosphodiesterase 7 (ENPP7) in both cohorts, with effect estimates ranging from −0.006 to −0.036. Significant concordant replicated associations were observed for delta-like canonical notch ligand (DLL1), inter-alpha-trypsin inhibitor heavy chain 3 (ITIH3), low-density lipoprotein receptor-related protein 11 (LRP11), regenerating family member 4 (REG4), and von Willebrand factor C domain containing protein 2 (VWC2) in both cohorts, with effect estimates ranging from 0.006 to 0.010. Effect estimates showed strong concordance between the two cohorts and overlapping confidence intervals for most proteins between cohorts. In UKB subgroup analysis, REG4 had significantly different effect sizes across European, African, and South Asian groups (p-het = 0.017) (Supplemental Figure 3). There were significant sex-specific differences in effect sizes for ITIH3, CREG1, and DXCR, in which there were large absolute effect sizes among females compared to males (p-het = 0.045, 0.033, and < 0.0001 respectively) (Supplemental Figure 4).

**Figure 2.**
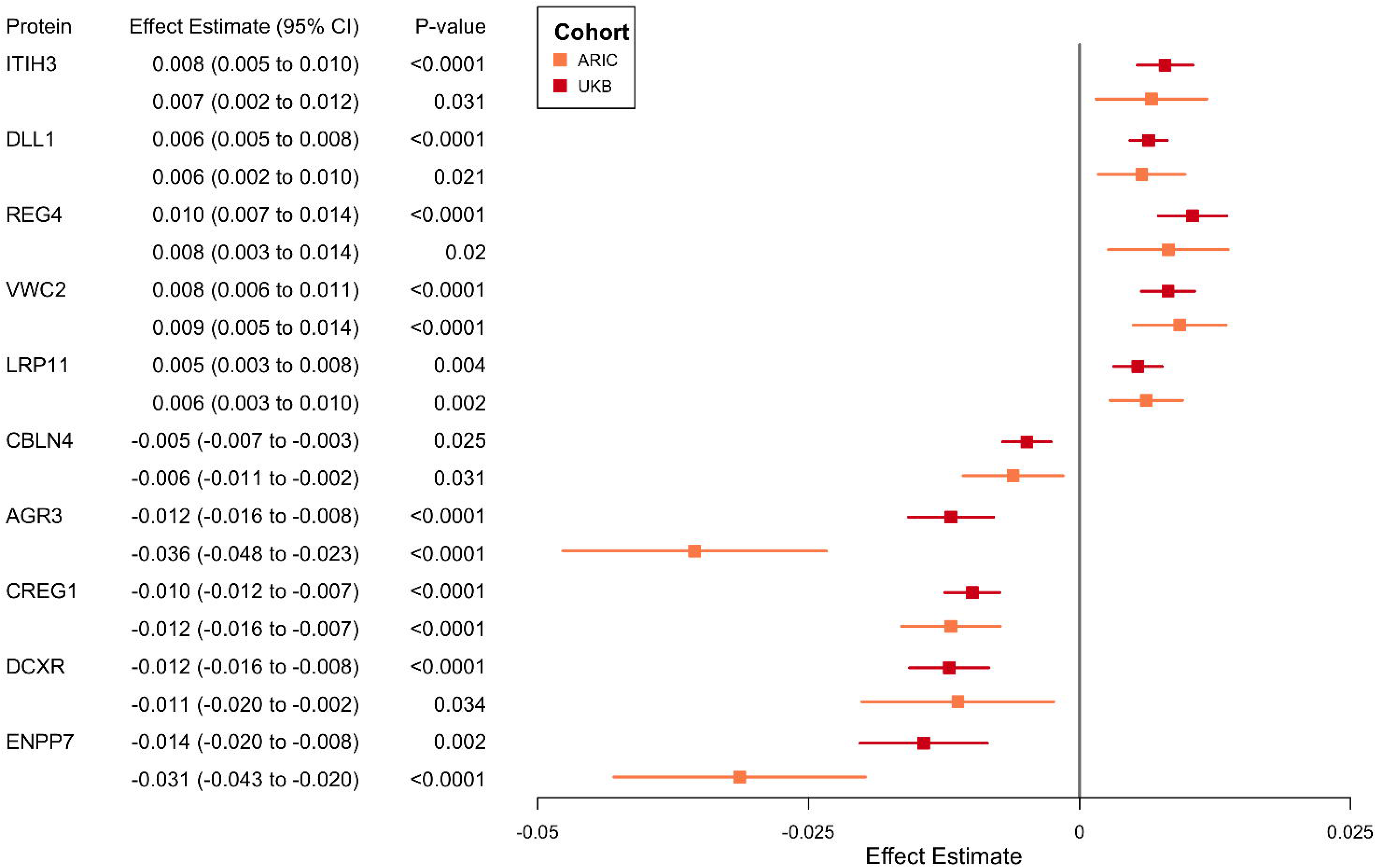
Association of Lp(a) per 50 nmol/L with circulating protein levels replicated in an independent cohort. Forest plot showing effect estimates (±95% CIs) for associations between Lp(a) per 50 nmol/L and circulating protein levels based on linear regression models adjusted for age at enrollment, sex, smoking, DM, BMI > 30, SBP, eGFR, LDL, HDL, TG, statin use, and PC1–PC10. Red indicates results from UKB and orange indicates results from ARIC. Multiple testing across proteins was addressed using the BH FDR correction, with FDR-adjusted p < 0.05 considered statistically significant. Abbreviations: ARIC, Atherosclerosis Risk in Communities; BMI, body mass index; CI, confidence interval; DM, diabetes mellitus; eGFR, estimated glomerular filtration rate; FDR, false discovery rate; HDL, high-density lipoprotein cholesterol; LDL, low-density lipoprotein cholesterol; Lp(a), lipoprotein(a); PC, principal component; SBP, systolic blood pressure; TG, triglycerides; UKB, United Kingdom Biobank.

### Secondary analyses

Given the high heritability of Lp(a), we assessed whether an *LPA* GRS was also correlated with Lp(a)-associated proteins (Supplemental Table 3). However, the *LPA* GRS was not significantly correlated with these 10 proteins (Figure 3). Since Lp(a) and LDL-C share highly similar morphology, we assessed protein associations with LDL-C adjusted for statin use per SD (mg/dL) (Supplemental Figure 5). Only two of the 10 proteins, REG4 (LDL-C p < 0.001) and VWC2 (LDL-C p < 0.001), had significant concordant associations with Lp(a) and LDL-C. Other proteins (AGR3, CBLN4, CREG1, DCXR, ENPP7, DLL1, and ITIH3) had significant inverse associations with Lp(a) and LDL-C. The only protein not associated with LDL-C was LRP11 (p = 0.233).

**Figure 3.**
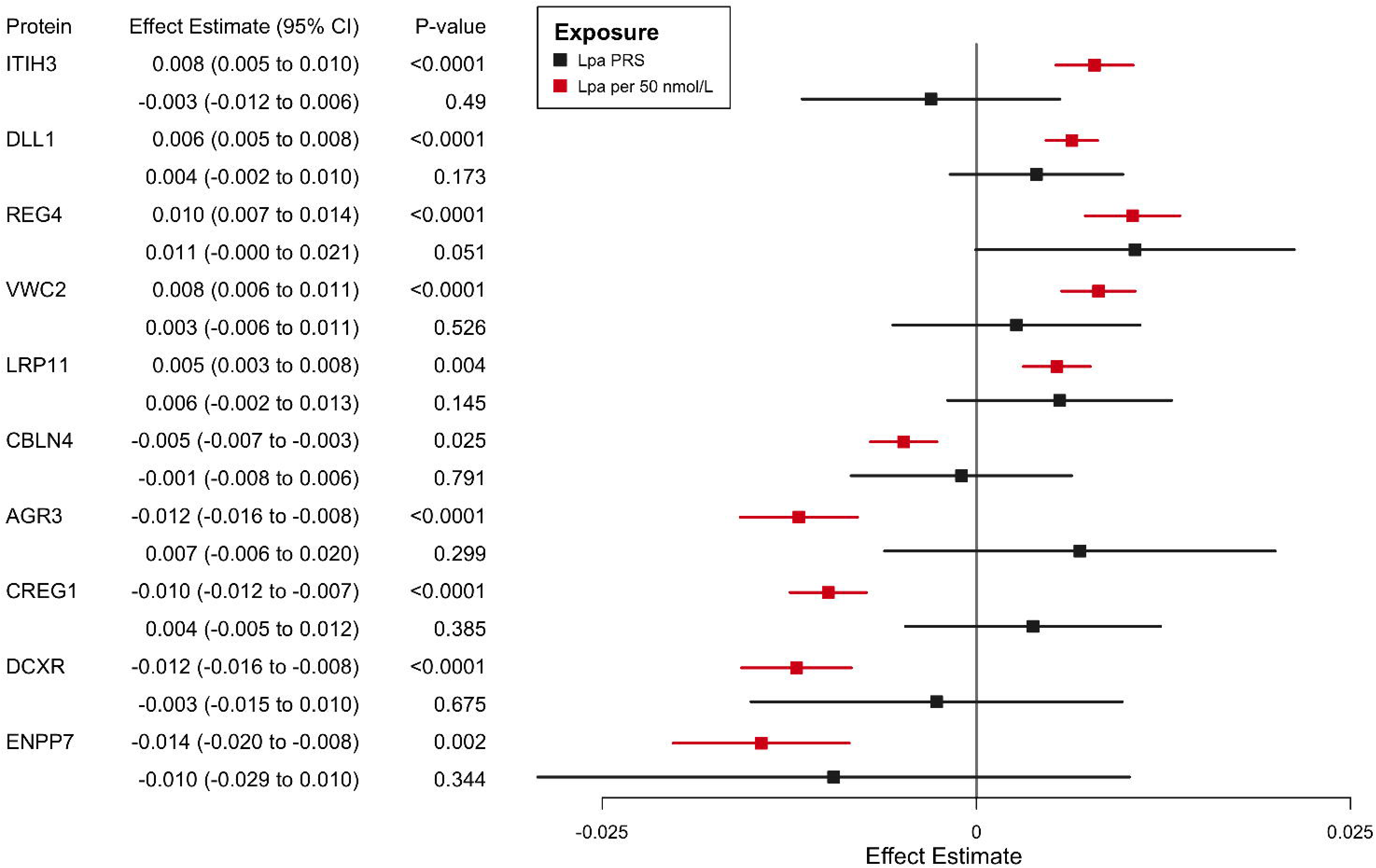
Circulating protein levels associated with Lp(a) per 50 nmol/L and an Lp(a) polygenic risk score. Forest plot showing effect estimates (±95% CIs) for associations between circulating protein levels and two exposures: the Lp(a) GRS (black squares) and measured Lp(a) per 50 nmol/L (red squares). Negative values indicate lower protein levels with higher Lp(a), while positive values indicate higher protein levels. Estimates were derived from linear regression models adjusted for age at enrollment, sex, smoking, DM, BMI > 30, SBP, eGFR, LDL, HDL, TG, and statin use, as well as PC1–PC10. Abbreviations: BMI, body mass index; CI, confidence interval; DM, diabetes mellitus; eGFR, estimated glomerular filtration rate; GRS, genetic risk score; HDL, high-density lipoprotein cholesterol; LDL, low-density lipoprotein cholesterol; Lp(a), lipoprotein(a); PC, principal component; SBP, systolic blood pressure; TG, triglycerides.

### Exploratory associations with ASCVD

We next examined the 10 proteins in the context of incident CAD, leveraging the largest ASCVD cohort available (Supplemental Table 4). Six of the 10 proteins showed directionally consistent associations with Lp(a) levels and dose-response relationships with incident CAD risk rates (Figure 4, Supplemental Figure 6, Supplemental Table 5). Specifically, proteins positively associated with Lp(a) (LRP11, DLL1, ITIH3, REG4, and VWC2) showed increasing CAD risk rates across increasing quartiles, while CBLN4, which was inversely associated with Lp(a), showed reduced CAD risk rates across increasing quartiles. Extending these analyses to other incident ASCVD and corresponding complications, there was a consistent, dose-dependent association with increasing concentrations of five proteins (LRP11, DLL1, ITIH3, REG4, and VWC2) (Supplemental Table 6).

**Figure 4.**
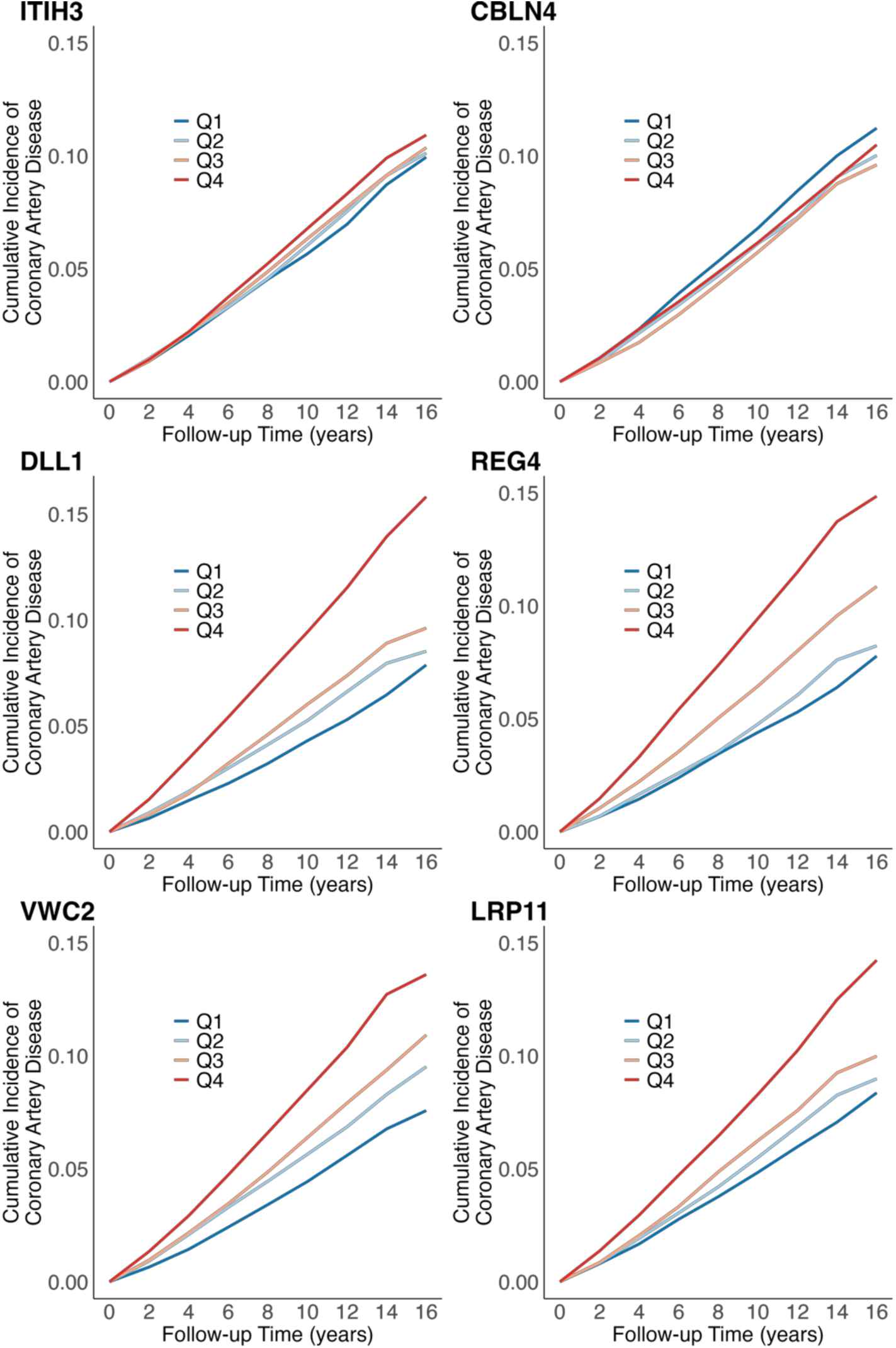
Cumulative incidence of CAD by quartiles of proteins directionally consistent with Lp(a). Kaplan–Meier curves showing the cumulative incidence of incident CAD across quartiles of circulating protein concentrations that were significantly associated with, and directionally consistent with, Lp(a). Participants were stratified into quartiles according to increasing protein levels. Proteins positively correlated with Lp(a) per 50 nmol/L were ITIH3, DLL1, LRP11, REG4, and VWC2. The protein negatively correlated with Lp(a) per 50 nmol/L was CBLN4. The expected direction of effect in relation to CAD risk was observed for all proteins (ITIH3, DLL1, LRP11, REG4, VWC2, and CBLN4). The absolute 10-year risk of CAD in this population was 6.20%. Abbreviations: CAD, coronary artery disease; Lp(a), lipoprotein(a); Q, quartile.

We subsequently examined associations between each protein and incident ASCVD using separate multivariable-adjusted Cox proportional hazards models for each ASCVD outcome. The proportional hazards assumption tested by Schoenfeld residuals was met by all outcomes assessed in this study (Supplemental Figure 7). Five proteins showed directionally consistent associations with Lp(a) and incident ASCVD (Figure 5, Supplemental Figure 8). Specifically, ITIH3 was significantly associated all ASCVD outcomes tested. For every one standard deviation increase in ITIH3, there relatively was a 13% increased risk of CAD (HR 1.13, 95% CI 1.04–1.23, p = 0.004), 42% increased risk of PAD (HR 1.42, 95% CI 1.19–1.69, p < 0.001), 65% increased risk of MALE (HR 1.65, 95% CI 1.14–2.40, p = 0.009), 45% increased risk of carotid stenosis (HR 1.45, 95% CI 1.13–1.85, p = 0.004), and 33% increased risk of ischemic stroke (HR 1.33, 95% CI 1.13–1.55, p < 0.001). DLL1 was significantly associated with only PAD (HR 1.61, 95% CI 1.26–2.05, p < 0.001). REG4 was significantly associated with CAD, PAD, MALE, and ischemic stroke. VWC2 was also significantly associated with PAD and MALE. CBLN4 was the only protein that had an inverse correlation with both Lp(a) and ASCVD, in which higher levels of CBLN4 were associated with a lower risk of CAD (HR 0.88, 95% CI 0.80–0.96, p=0.007), PAD (HR 0.78, 95% CI 0.64–0.96, p=0.016), and ischemic stroke (HR 0.72, 95% CI 0.60–0.85, p < 0.001). Time-varying analyses showed that while the strength of associations diminished over time, the associations remained significant across all life years evaluated (Supplemental Figure 9).

**Figure 5.**
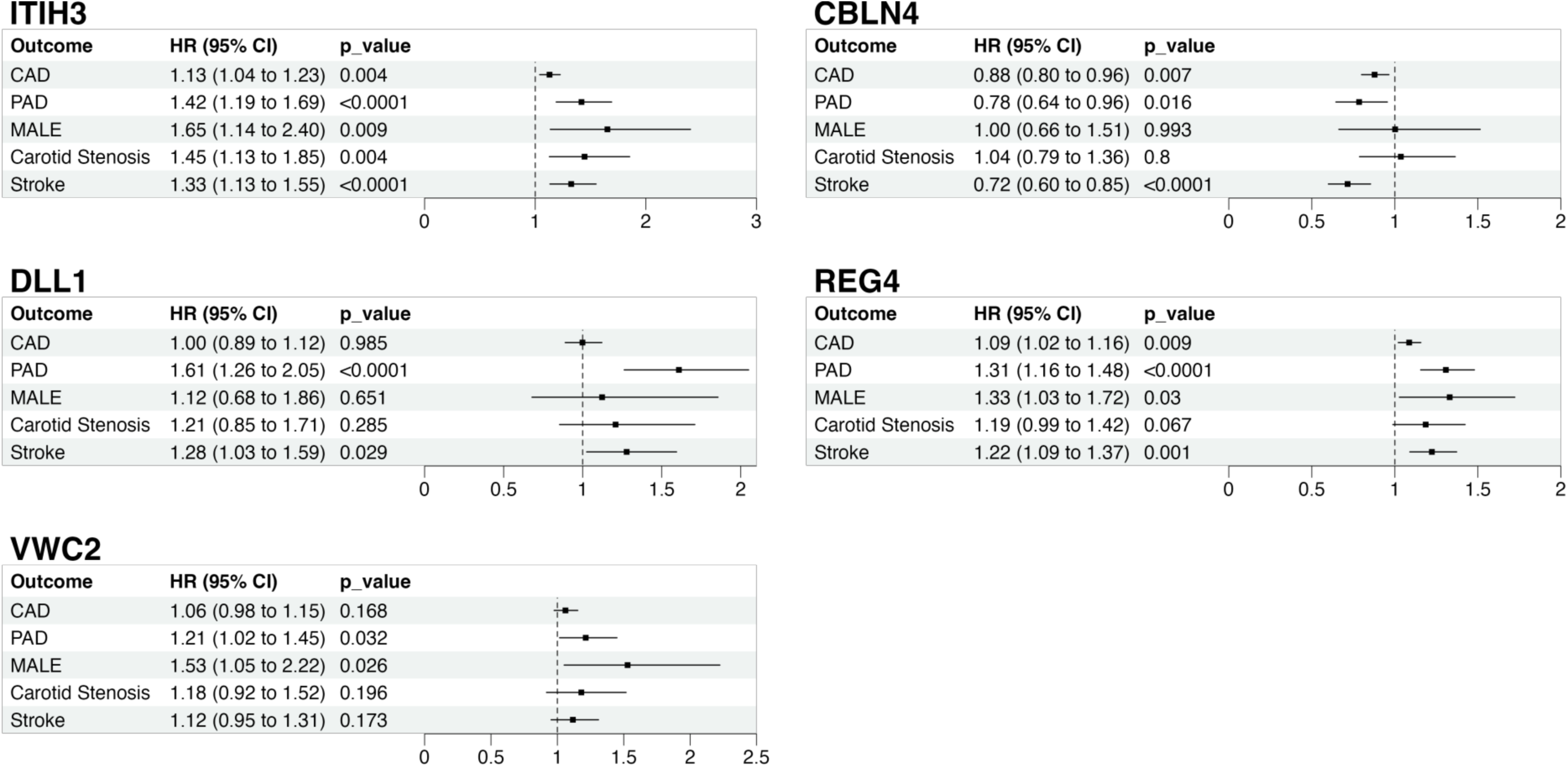
Risk of incident ASCVD per circulating protein level directionally consistent with Lp(a). HRs and corresponding 95% CIs for incident ASCVD are shown per unit increase in circulating protein levels. Each row represents a separate Cox proportional hazards model adjusted for age at enrollment, sex, smoking, DM, BMI > 30, SBP, eGFR, LDL, HDL, TG, statin use, and PC1–PC10. Proteins positively correlated with Lp(a) per 50 nmol/L were ITIH3, DLL1, REG4, and VWC2. The protein negatively correlated with Lp(a) per 50 nmol/L was CBLN4. The expected direction of effect in relation to ASCVD risk was observed for all proteins shown. Abbreviations: ASCVD, atherosclerotic cardiovascular disease; BMI, body mass index; CI, confidence interval; DM, diabetes mellitus; eGFR, estimated glomerular filtration rate; HDL, high-density lipoprotein cholesterol; HR, hazard ratio; LDL, low-density lipoprotein cholesterol; Lp(a), lipoprotein(a); PC, principal component; SBP, systolic blood pressure; TG, triglycerides.

## DISCUSSION

In this study, we identified plasma proteins correlated with Lp(a) that were also associated with ASCVD clinical outcomes. In a discovery analysis comprising over 40,000 participants, we identified 164 proteins associated with Lp(a) levels. Of the 164 proteins, 10 replicated in an external cohort. Despite the high heritability of Lp(a), the genetic drivers of elevated *LPA* were not correlated with associated proteins. Finally, five of these proteins were associated with incident ASCVD. These findings provide insights into new cellular and molecular pathways that may link Lp(a) to ASCVD.

The presence of replicated proteomic associations with the Lp(a) biomarker but not with *LPA* GRS advances new hypotheses. Whether Lp(a) is the cause, consequence, or correlated with other responsible factors merits further investigation. Nevertheless, many correlated factors are associated with incident ASCVD. While Lp(a) is generally stable across the life course due to high heritability, those with the highest concentrations have the highest degree of variability.^24^ Lp(a) is an acute phase reactant potentially responsive to key inflammatory proteins identified.^34^ Hormonal changes, including exogeneous estrogen and pregnancy, are also correlated with Lp(a) levels.^35^ As enumerated in further detail, the mechanisms by which identified factors are linked to Lp(a)-associated ASCVD require further study.

Our analyses identified several inflammatory proteins strongly associated with Lp(a) and incident ASCVD, distinct from LDL-C. Previous research in a small cohort of healthy human volunteers demonstrated that ITIH3 was more abundant on Lp(a) compared to LDL-C in circulation.^36^ Similarly, we observe that Lp(a) is positively correlated with ITIH3, a protein involved in cell adhesion that may enhance leukocyte migration and inflammation.^37^ ITIH3 is synthesized in the liver as light- and heavy-chain polypeptides, which together constitute the majority of circulating ITIH3.^38^ However, ITIH3 expression and function in circulation may differ at the tissue-level. For example, RNA-seq data from 262 human liver biopsies demonstrated a negative correlation between hepatic ITIH3 expression and non-alcoholic fatty liver disease (NAFLD) phenotypes.^39^ In our study, ITIH3 was significantly associated with incident ASCVD, including CAD, PAD, MALE, carotid stenosis, and ischemic stroke, but tissue-level effects may vary.^40^

The strong association between Lp(a) and vascular calcification suggests a shared osteogenic signaling pathway through which Lp(a)-related proteins, such as DLL1 and VWC2, may promote atherosclerosis and calcific disease. In our study, DLL1 (Delta-like protein 1) was associated with both incident PAD and ischemic stroke. DLL1 is a ligand in the Notch signaling pathway contributing to inflammatory processes that lead to osteogenic differentiation.^41^ A study comparing 136 patients with symptomatic calcific aortic stenosis had higher levels of serum DLL1 compared to 95 healthy human controls, suggesting that Notch signaling may cause osteogenic differentiation resulting in calcification of the aortic valve.^42^ Indeed, Lp(a) is a recognized potentially causal factor for calcific aortic valve stenosis.^5^ VWC2 is also involved in osteogenesis by inhibiting the Activin–Smad2 signaling pathway^43^ and antagonizing several bone morphogenetic proteins^44^ that exert protective effects against vascular calcification.^45^ Recent single-cell transcriptomic and spatial profiling studies have shown that VWC2 is expressed in human vascular cells and arterial tissue,^46^ consistent with our findings that VWC2 is associated with incident PAD and MALE.

Other Lp(a)-associated proteins identified in this study have known links to lipid metabolism, such as CBLN4, CREG1, REG4, ENPP7, and LRP11. CBLN4 was negatively associated with Lp(a) as well as CAD, PAD, and ischemic stroke. Although CBLN4 has primarily been studied in the brain and spinal cord,^47^ it has also been implicated in lipid particle uptake.^48^ CREG1 was inversely associated with Lp(a), consistent with previous reports that ox-LDL-C directly downregulates CREG1 in vascular cells via epigenetic mechanisms^49^ and may confer a protective effect against atherosclerosis.^50^ REG4 also has a distinct link to lipid metabolism. In a study of 258 patients from a Chinese cohort with metabolic syndrome, circulating REG4 levels were significantly higher compared to healthy controls, and LDL-C cholesterol was found to independently influence serum REG4.^51^ Similarly, our study showed a consistent association between REG4 and LDL-C cholesterol in a predominantly European cohort. ENPP7 is a phospholipase with a direct role in intestinal lipid absorption.^52^ LRP11 (Low-density lipoprotein receptor-related protein 11), although initially studied in hepatocellular carcinoma,^53^ has emerging evidence supporting its role in lipid metabolism.^54^

### Limitations

This observational study, conducted within a large biobank, should be interpreted in the context of the inherent limitations of such cohorts, including issues related to population representativeness, measurement error, and the observational nature of association analyses. First, the study population was predominantly of European ancestry, limiting the generalizability of our findings. Second, protein measurements obtained via the Olink platform were collected at enrollment and may also be subject to assay variability. However, our replication strategy used a distinct proteomic platform, but this may have also reduced power for replication. Third, given the observational nature of the study, directionality and causality of associations may not be suitably claimed.

## CONCLUSIONS

While Lp(a) has been extensively characterized structurally and epidemiologically, the cellular and molecular mechanisms by which it contributes to ASCVD remain incompletely understood. In this study, we leveraged high-throughput proteomic profiling to examine the relationship between circulating Lp(a) and protein expression, and to evaluate the association of these proteins with incident ASCVD events. We identified 10 replicated protein candidates linked to Lp(a), several of which appear to act independently of genetic Lp(a) risk and distinctly from LDL-C. These findings provide a foundation for future mechanistic studies aimed at elucidating the roles of these proteins in Lp(a)-mediated atherogenesis and may inform the development of protein-targeted strategies for the prevention of ASCVD.

## Abbreviations

Lp(a): Lipoprotein(a)
UKB: United Kingdom Biobank
ARIC: Atherosclerosis Risk in Communities
SD: Standard Deviation
IQR: Interquartile Range
LDL-C: Low-density Lipoprotein Cholesterol
HDL-C: High-density Lipoprotein Cholesterol
HR: Hazard Ratio
KNN: K-nearest Neighbors
CKD: Chronic Kidney Disease

## Data Availability

The data that support the findings of this study are available from the UK Biobank upon reasonable request.

https://biobank.ndph.ox.ac.uk/

## ACKNOWLEDGEMENTS

The data that support the findings of this study are available from the UK Biobank upon reasonable request. Code to perform analyses in this manuscript are available from the authors upon request. The Atherosclerosis Risk in Communities study has been funded in whole or in part with Federal funds from the National Heart, Lung, and Blood Institute, National Institutes of Health, Department of Health, and Human Services, under Contract nos. (75N92022D00001, 75N92022D00002, 75N92022D00003, 75N92022D00004, 75N92022D00005). The authors thank the staff and participants of the ARIC study for their important contributions.

