## Supplemental Figures 1-9 for "Proteomic analysis identifies lipoprotein(a)-associated proteins linked to incident atherosclerotic cardiovascular disease events"

**Table of Contents**

**Supplemental Tables:**

Supplemental Table 1. Association of lipoprotein(a) per 50 nmol/L with the outcome of continuous protein levels in the UK Biobank.

Supplemental Table 2. Association of lipoprotein(a) per 50 nmol/L with continuous protein levels in the Atherosclerosis Risk in Communities.

Supplemental Table 3. Association between lipoprotein(a) and other known biomarkers including a polygenic risk score and low density-lipoprotein.

Supplemental Table 4. Atherosclerotic cardiovascular disease incidence and prevalence in the UK Biobank cohort.

Supplemental Table 5. Cumulative incidence of coronary artery disease by protein expression quartiles over 16 years of follow-up.

Supplemental Table 6. Cumulative incidence of atherosclerotic cardiovascular disease by protein expression quartiles over 16 years of follow-up.

**Supplemental Figures:**

Supplemental Figure 1. Flowchart of inclusion and exclusion criteria in UK Biobank	and Atherosclerosis Risk in Communities.

Supplemental Figure 2. Proteins significantly associated with lipoprotein(a) per 50	nmol/L annotated using the UniProt in purple and Gene Ontology in green.

Supplemental Figure 3. Association of lipoprotein(a) per 50 nmol/L with circulating	protein levels stratified by ancestry.

Supplemental Figure 4. Association of lipoprotein(a) per 50 nmol/L with circulating	protein levels stratified by sex.

Supplemental Figure 5. Circulating protein levels associated with lipoprotein(a) and low-	density lipoprotein cholesterol.

Supplemental Figure 6. Proportional hazards assumption test by each protein.

Supplemental Figure 7. Risk of incident atherosclerotic disease per circulating protein	level directionally inconsistent with lipoprotein(a).

Supplemental Figure 8. Dynamic hazard ratio of atherosclerotic disease for proteins by	age at Enrollment.

Supplemental Figure 9. Dynamic HRs for ASCVD by protein level and age at	enrollment.

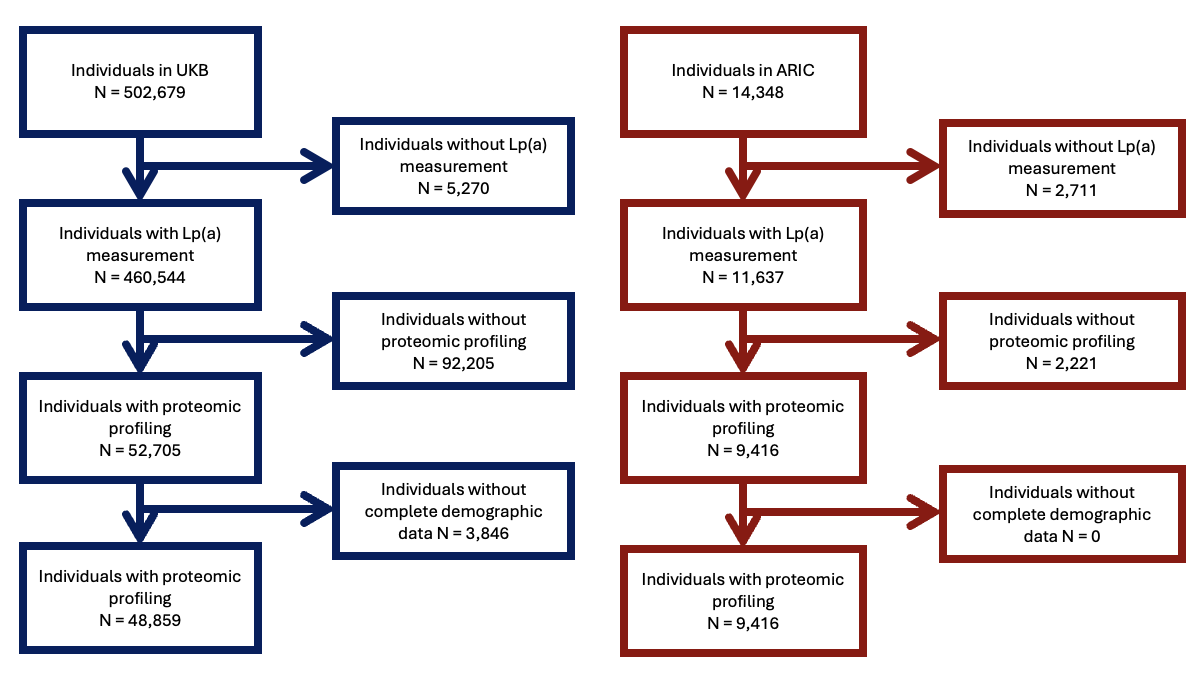

**Supplemental Figure 1.** Flowchart of inclusion and exclusion criteria in UKB and ARIC. Abbreviations: ARIC, Atherosclerosis Risk in Communities; UKB, United Kingdom Biobank.

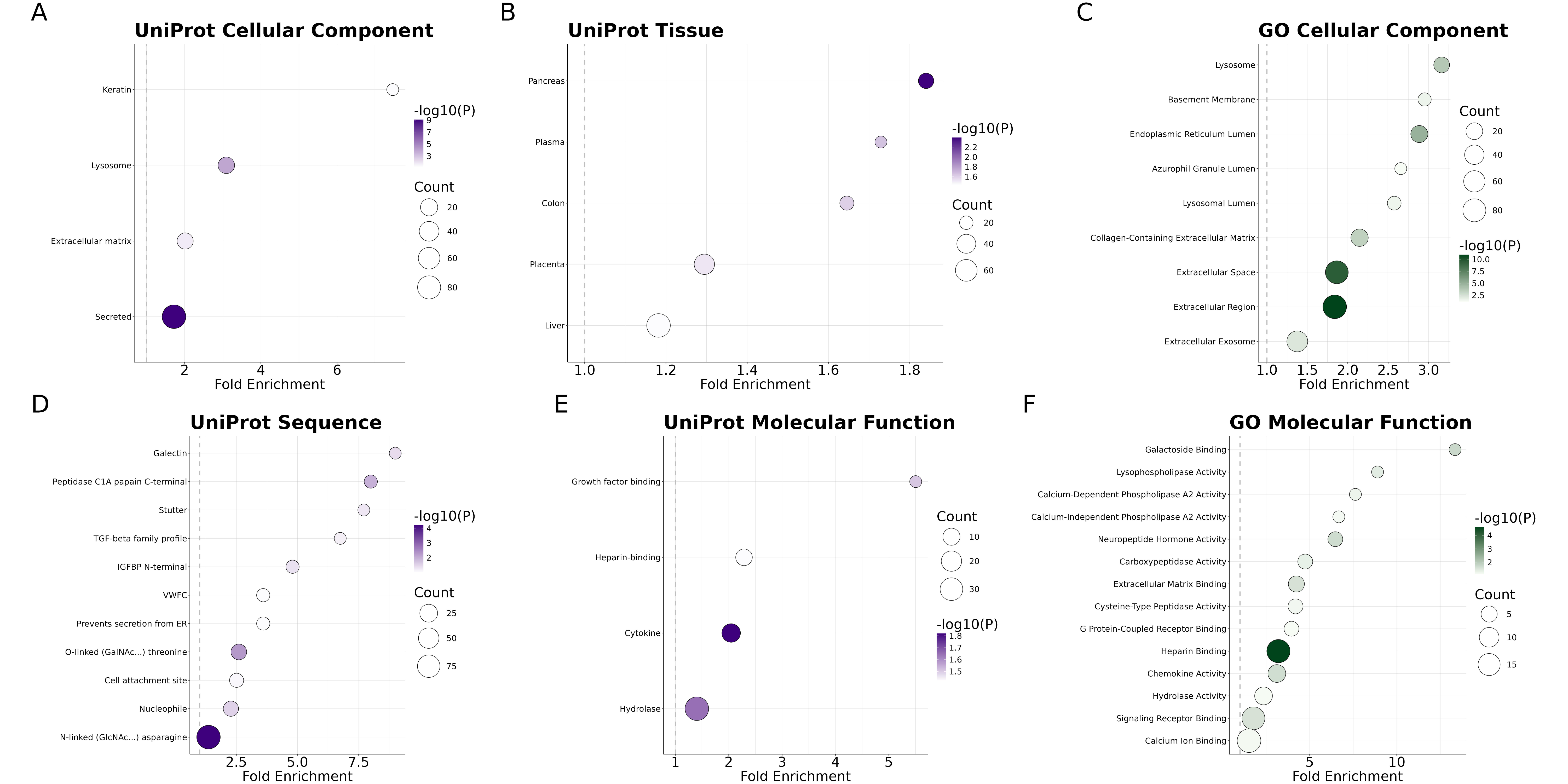

**Supplemental Figure 2.** Proteins significantly associated with Lp(a) per 50 nmol/L annotated using UniProt (purple) and GO (green). UniProt annotation schemes included cellular components, molecular function, sequence features, and tissue categories. GO annotation schemes included cellular components and molecular functions. The darkness of each point reflects statistical significance, with darker points indicating larger −log10(p) values. Point size represents the number of proteins annotated within each category, with larger circles indicating a greater number of proteins. Abbreviations: GO, Gene Ontology; Lp(a), lipoprotein(a).

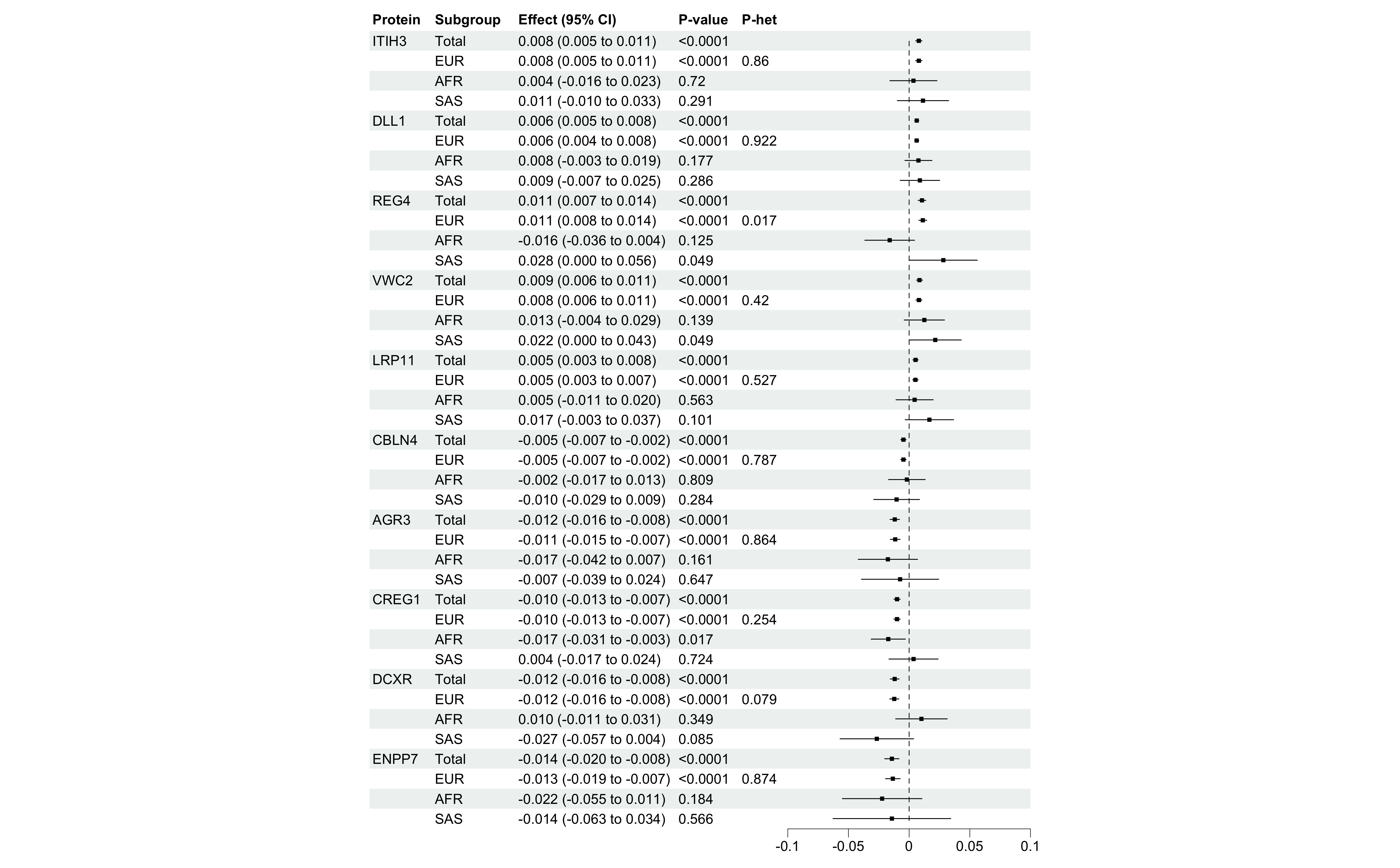

**Supplemental Figure 3.** Association of Lp(a) per 50 nmol/L with circulating protein levels stratified by ancestry. Effect estimates with corresponding 95% CIs were obtained from linear regression models adjusted for age at enrollment, sex, smoking, DM, BMI > 30, SBP, eGFR, LDL, HDL, TG, statin use, and PC1–PC10. Abbreviations: BMI, body mass index; CI, confidence interval; DM, diabetes mellitus; eGFR, estimated glomerular filtration rate; HDL, high-density lipoprotein cholesterol; LDL, low-density lipoprotein cholesterol; Lp(a), lipoprotein(a); PC, principal component; SBP, systolic blood pressure; TG, triglycerides.

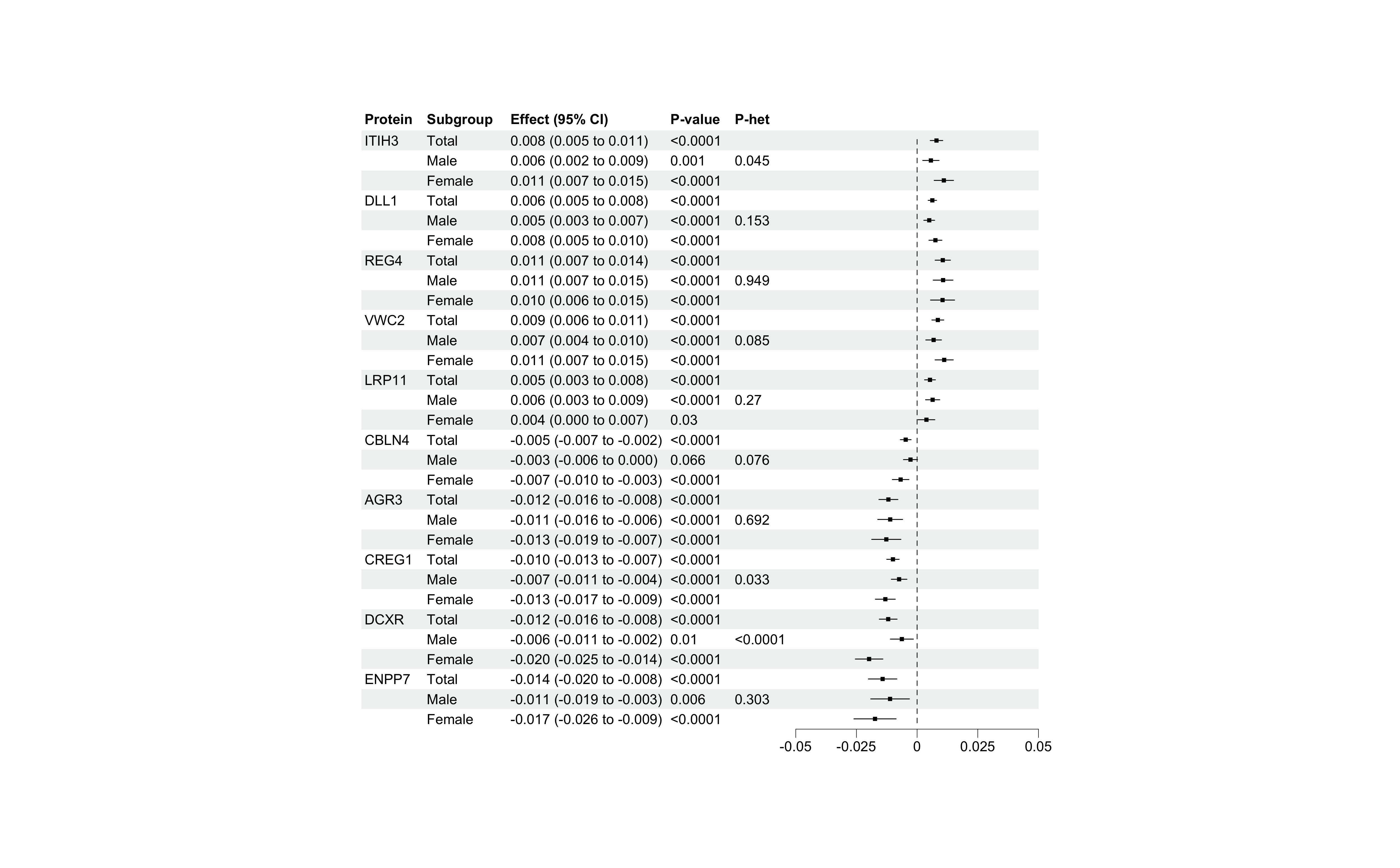

**Supplemental Figure 4.** Association of Lp(a) per 50 nmol/L with circulating protein levels stratified by sex. Effect estimates with corresponding 95% CIs were obtained from linear regression models adjusted for age at enrollment, smoking, DM, BMI > 30, SBP, eGFR, LDL, HDL, TG, statin use, and PC1–PC10. Abbreviations: BMI, body mass index; CI, confidence interval; DM, diabetes mellitus; eGFR, estimated glomerular filtration rate; HDL, high-density lipoprotein cholesterol; LDL, low-density lipoprotein cholesterol; Lp(a), lipoprotein(a); PC, principal component; SBP, systolic blood pressure; TG, triglycerides.

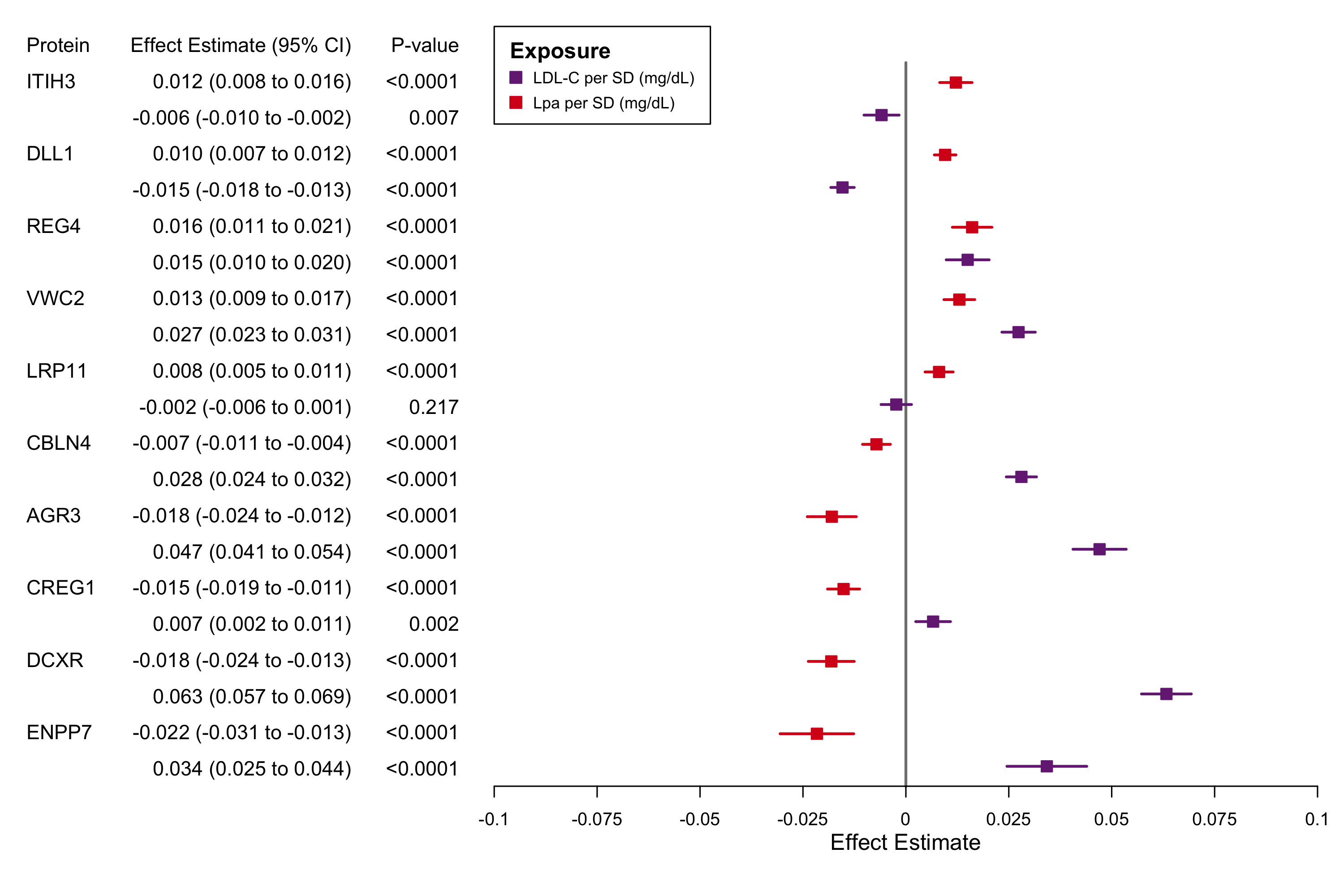

**Supplemental Figure 5.** Circulating protein levels associated with Lp(a) and LDL. Forest plot showing effect estimates (±95% CIs) for associations between circulating protein levels and two exposures: Lp(a) per SD (mg/dL) and LDL adjusted for statin use. Estimates were derived from linear regression models adjusted for age at enrollment, smoking, DM, BMI > 30, SBP, eGFR, HDL, TG, and PC1–PC10. Abbreviations: BMI, body mass index; CI, confidence interval; DM, diabetes mellitus; eGFR, estimated glomerular filtration rate; HDL, high-density lipoprotein cholesterol; LDL, low-density lipoprotein cholesterol; Lp(a), lipoprotein(a); PC, principal component; SBP, systolic blood pressure; SD, standard deviation; TG, triglycerides.

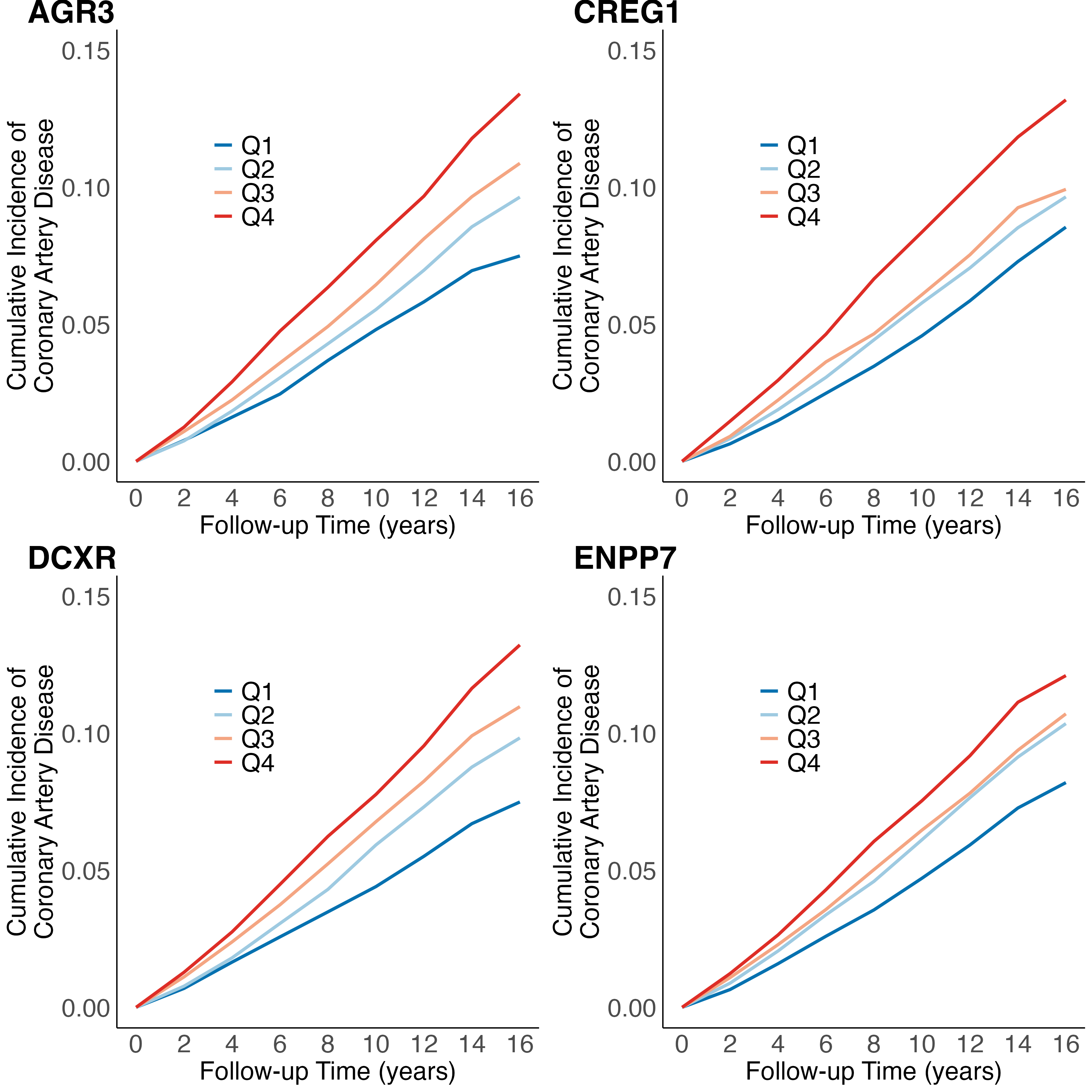

**Supplemental Figure 6.** Cumulative incidence of CAD by quartiles of proteins directionally inconsistent with Lp(a). Kaplan–Meier curves showing the cumulative incidence of incident CAD across quartiles of circulating protein concentrations significantly associated with, but directionally inconsistent with, Lp(a). Participants were stratified into quartiles according to increasing protein levels. Proteins negatively correlated with Lp(a) per 50 nmol/L were AGR3, CREG1, DCXR, and ENPP7, yet demonstrated increasing CAD risk across higher quartiles. The absolute 10-year risk of CAD in this population was 6.20%. Abbreviations: CAD, coronary artery disease; Lp(a), lipoprotein(a); Q, quartile.

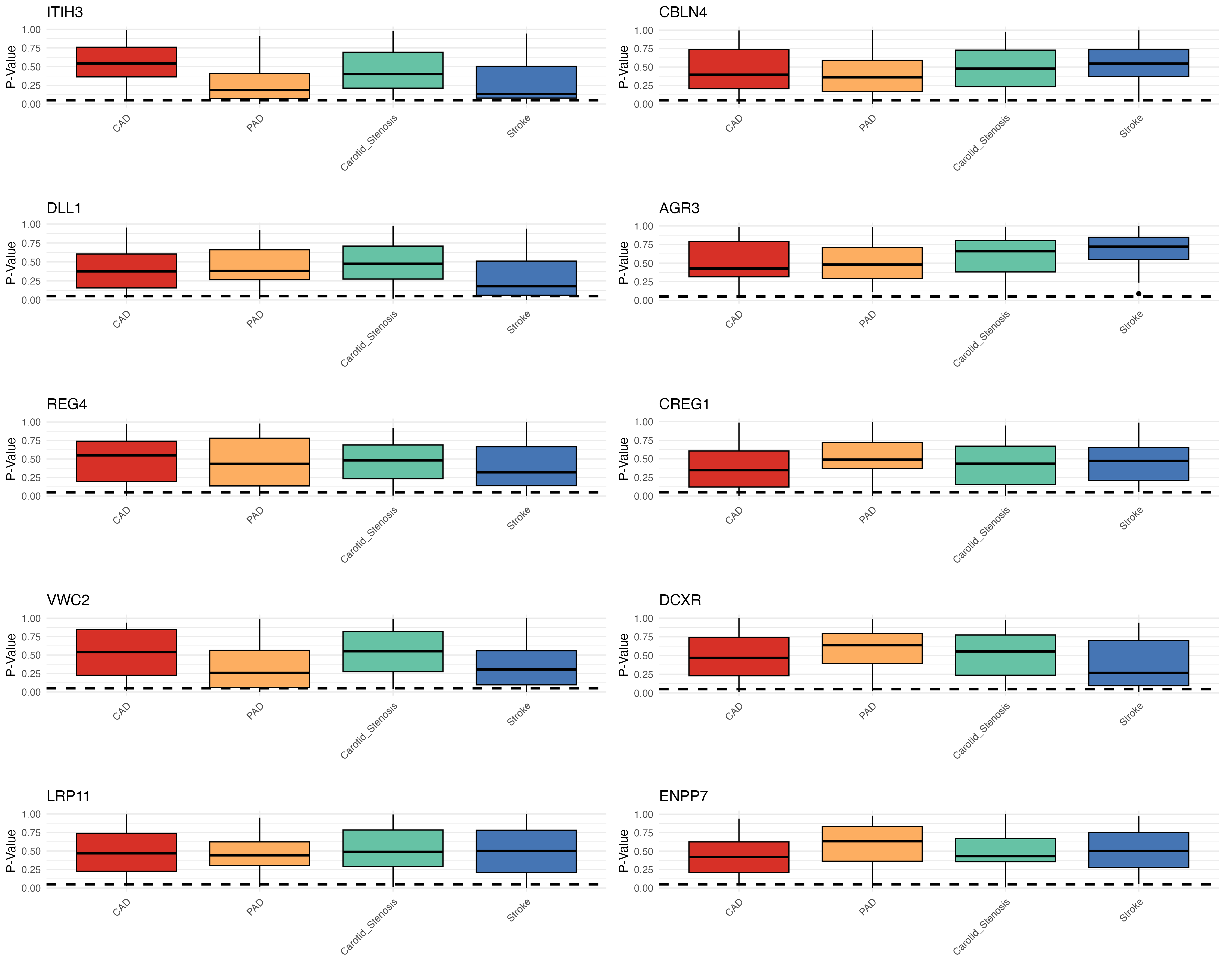

**Supplemental Figure 7.** Proportional hazards assumption test for each protein. Scaled Schoenfeld residuals were tested for correlation with time using Cox proportional hazards models to assess the proportionality assumption at 1-year increments. Values were averaged over 10 years to generate median and IQRs. Proteins positively correlated with Lp(a) per 50 nmol/L in the first column were ITIH3, DLL1, LRP11, REG4, and VWC2. Proteins negatively correlated with Lp(a) per 50 nmol/L in the second column were AGR3, CBLN4, CREG1, DCXR, and ENPP7. Abbreviations: IQR, interquartile range; Lp(a), lipoprotein(a).

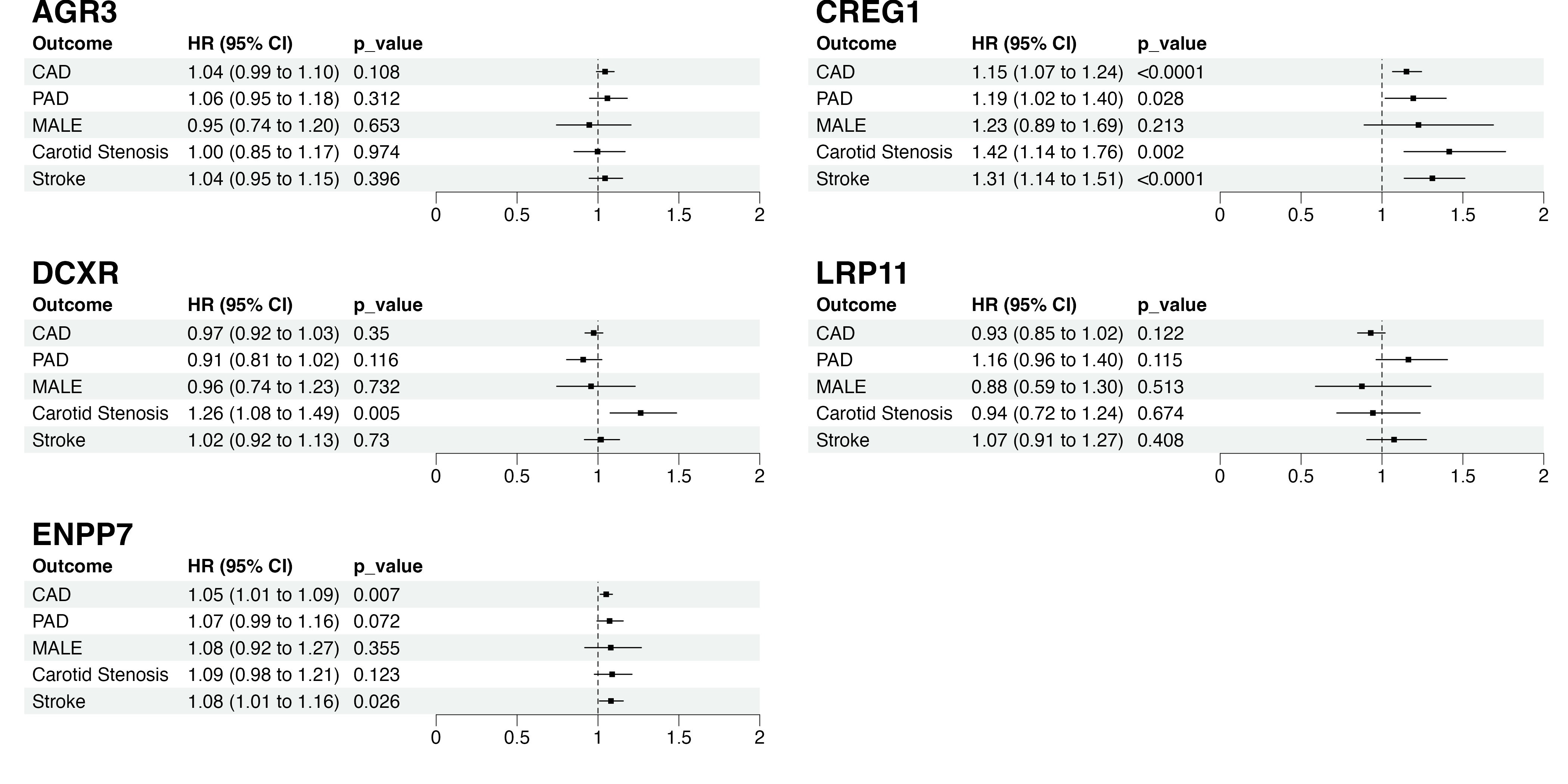

**Supplemental Figure 8.** Risk of incident ASCVD per circulating protein level directionally inconsistent with Lp(a). HRs and corresponding 95% CIs for incident ASCVD are shown per unit increase in circulating protein levels. Each row represents a separate Cox proportional hazards model adjusted for age at enrollment, sex, smoking, DM, BMI > 30, SBP, eGFR, LDL, HDL, TG, statin use, and PC1–PC10. The protein positively correlated with Lp(a) per 50 nmol/L was LRP11. Proteins negatively correlated with Lp(a) per 50 nmol/L were AGR3, CREG1, DCXR, and ENPP7. Abbreviations: ASCVD, atherosclerotic cardiovascular disease; BMI, body mass index; CI, confidence interval; DM, diabetes mellitus; eGFR, estimated glomerular filtration rate; HDL, high-density lipoprotein cholesterol; HR, hazard ratio; LDL, low-density lipoprotein cholesterol; Lp(a), lipoprotein(a); PC, principal component; SBP, systolic blood pressure; TG, triglycerides.

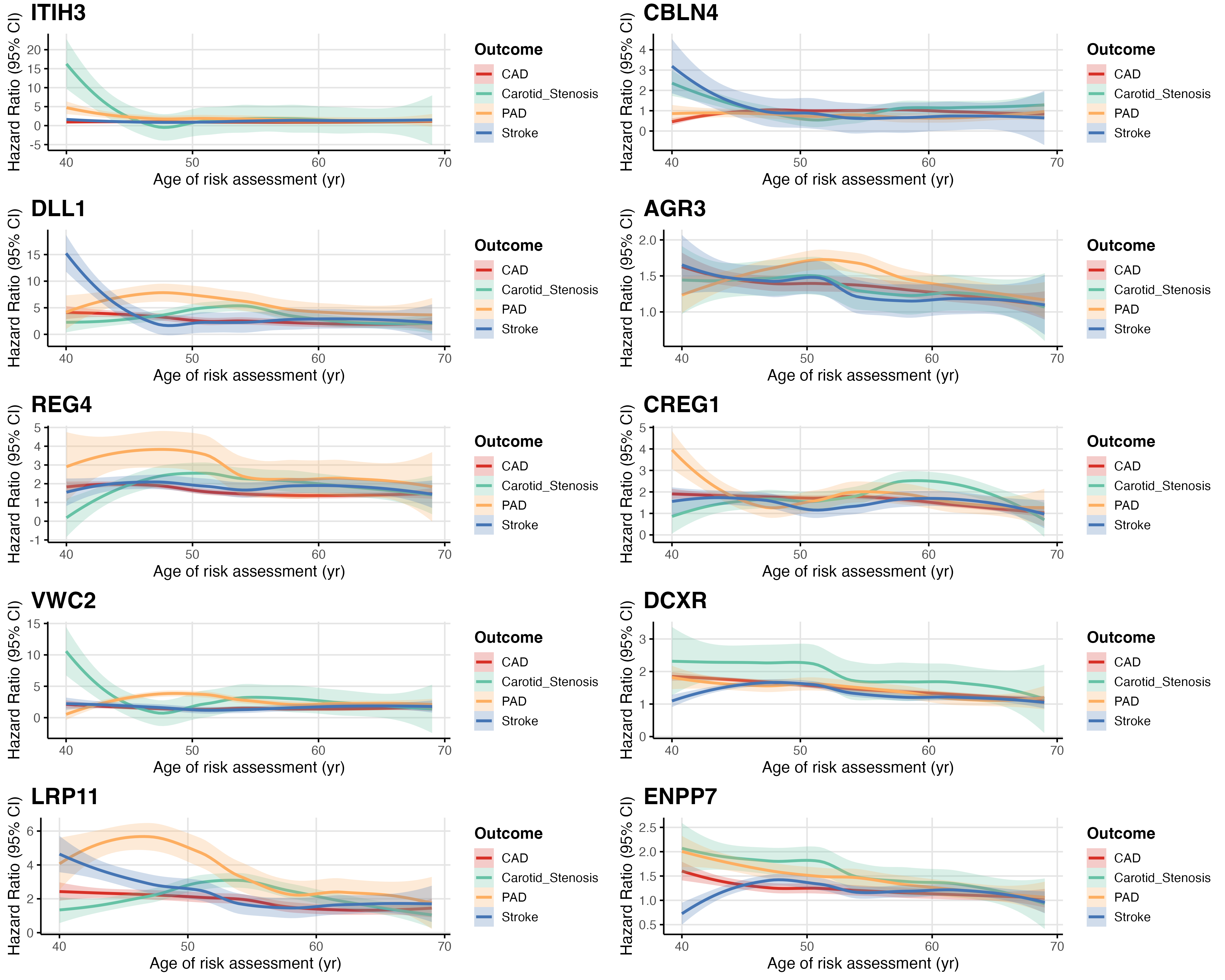

**Supplemental Figure 9.** Dynamic HRs for ASCVD by protein level and age at enrollment. Age-specific HRs for incident ASCVD are plotted for each protein across ages at enrollment. HRs were obtained from Cox proportional hazards models using each ASCVD subtype as a separate outcome, where red represents CAD, green represents carotid stenosis, orange represents PAD, and blue represents ischemic stroke. No covariates were included to isolate the effect of each protein. Each panel represents a different protein. Proteins positively correlated with Lp(a) per 50 nmol/L in the first column were ITIH3, DLL1, LRP11, REG4, and VWC2. Proteins negatively correlated with Lp(a) per 50 nmol/L in the second column were AGR3, CBLN4, CREG1, DCXR, and ENPP7. Abbreviations: ASCVD, atherosclerotic cardiovascular disease; CAD, coronary artery disease; HR, hazard ratio; Lp(a), lipoprotein(a); PAD, peripheral artery disease.
